# Post-tuberculosis sequelae and their correlation with quality of life: An observational study at a tertiary care center of north India

**DOI:** 10.1101/2021.10.04.21264524

**Authors:** Nishant Aggarwal, Tamoghna Ghosh, Munish Bhan, Vignesh Dwarakanathan, Prayas Sethi, Ved Prakash Meena, Sanjeev Sinha, Animesh Ray

## Abstract

**Background:** Lung impairment is a frequently recognised outcome in patients treated for pulmonary tuberculosis (TB). The impact of post-TB sequelae is not only restricted to clinical outcomes but also includes the quality of life and psycho-social well-being. However the magnitude of involvement of quality of life and the likely factors determining it are not clear. In this study, we assess the degree of compromise of quality of life and its determinants in patients of post-TB sequelae.

**Methods:** Patients >18 years of age with a history of pulmonary tuberculosis were included in the study. Clinical history, pulmonary function test (PFT) and chest radiographs were recorded. The severity of dyspnea was evaluated using mMRC; quality of life assessment (QoL) was done using two standardized questionnaires- St. George’s Respiratory Questionnaire (SGRQ) and Seattle Obstructive Lung Disease Questionnaire (SOLQ).

**Results:** A total of 90 participants (mean age 40.4±11.6 years; 60 [66.7%] males) were recruited in the study. Overall, 93.3% (95% CI: 86.1%-97.5%) were currently symptomatic. A total of 96.8% (61/63) patients had abnormal pulmonary function test results, out of which 61.9% (39/63) had a mixed type. Out of 65 patients with chest radiographs available, 60 (92.3%,95% CI:84.6-96.8) patients had abnormalities with 45 (69.2%,95% CI: 58.3-78.2) having bilateral chest radiographic abnormalities. On assessment of QoL by SGRQ, the average score obtained was 42.3±24.0 (95% CI:37.3-47.3), with ‘Symptoms’ being the most affected domain. On using SOLQ, ‘Treatment satisfaction’ (mean score 38.5±21.7, 95% CI:34-43) and ‘Physical function’ (mean score 66.6±23.7, 95% CI:61.6-71.6) were the most affected. Neither spirometry results nor chest radiograph severity score could significantly account for the reduction of QoL. There was also a significant correlation between most of the parameters of SGRQ and SOLQ questionnaires in these patients.

**Conclusion:** Persistent respiratory symptoms, radiographic and spirometric abnormalities, were present in most of the patients with post-tubercular sequelae. Quality of life, as measured by SGRQ and SOLQ, was found to be significantly impaired in the majority of patients.

## 1. INTRODUCTION

Post-tubercular lung disease is defined as ‘Evidence of chronic respiratory abnormality, with or without symptoms, attributable at least in part to previous tuberculosis.’^1^ An estimated 18-87% of the patients with pulmonary tuberculosis (TB) experience lung impairment^2^ and may have mortality rate of upto three times that of general population.^3^ The actual burden of post-TB sequelae may be underreported in developing countries where clinical services, research and advocacy are far from adequate. In addition, it is hard to fully measure the impact of these sequelae as they affect not just the individual, but also their families and entire communities.^1^

Even after successful treatment, many patients suffer from post-TB sequelae. These may be structural complications (such as bronchiectasis, broncholithiasis, residual cavitation, chronic obstructive pulmonary disease [COPD]), infectious complications (such as COPD exacerbations, *Aspergillus fumigatus* infections, non-tubercular mycobacterial infections, pneumonia), or psychosocial morbidities (such as anxiety, depression, financial burden).^4^

Tuberculosis (TB) disease causes detrimental impact on the Health Related Quality of Life (HRQoL) of the patient.^4^ The need for evaluation of therapeutic interventions for TB is no longer restricted to clinical outcomes but also includes psycho-social well-being and HRQoL. The measurements of HRQoL are being used in direct patient care processes, clinical trials, program evaluations, and for monitoring health status in populations. Quality of life has become a very important outcome measure in clinical research, and advances have been made in assessing the impact of many diseases on HRQoL. There is a lack of quantitative evidence on the clinical features in patients with long term post-TB sequelae and their impact on the health-related quality of life (QoL). Therefore, the objectives of our study were to describe the symptomatic profile and radiological features in patients with post-TB sequelae. We also aim to describe the QoL in these patients and compare it to the clinical symptoms and correlate the scores obtained in SGRQ and SOLQ.

## 2. METHODS

This was a cross-sectional single center study performed in the Chest Clinic of a tertiary care academic medical center in northern India from October 2019 to September 2020. The study was reported using the Strengthening the Reporting of Observational Studies in Epidemiology (STROBE) guidelines.^5^ Patients >18 years of age who had a history of pulmonary tuberculosis (who had received treatment on the basis of microbiological reports or on clinician’s discretion) whose current symptoms could not be explained by a concomitant disease were included in the study. Children, pregnant women, or patients with another coexisting pulmonary disease were not included in the study. The study protocol was approved by the institute ethics committee (IECPG-652/28.11.2019) and conforms to the ethical guidelines of the 1975 Declaration of Helsinki as reflected in a prior approval by the institution’s human research committee.

A detailed history was elicited by NA in one-on-one interviews with the patients on a pre-designed proforma, which included demographic details, clinical symptoms and their severity, and the use of inhalers. Height (in cm) and weight (in kg) were measured in the clinic and the body mass index (BMI) was calculated. Modified kuppuswamy scale was used to assess socio-economic status.^6^ The severity of dyspnea was evaluated using the modified Medical Research Council scale (mMRC). Pulmonary function test (PFT) parameters (forced vital capacity [FVC], forced expiratory volume in 1 second [FEV1], and FEV1/FVC) were also noted along with their percentage predicted value. Obstructive disease was defined as FEV1/FVC ratio <0.8, with a FVC ≥80% of the predicted value. Restrictive disease was diagnosed as FEV1/FVC ratio >0.8 and FVC was <80% of the predicted values. Combined disease was defined as an FEV1/FVC ratio <0.8 and FVC <80% of the predicted value.^7,8^ The severity of abnormalities on chest radiographs was scored from 0-5 for the four quadrants (right upper, right lower, left upper, left lower quadrant) according to the scoring system developed by Báez-Saldaña et al.^9^

The proforma also included two QoL questionnaires- the St. George’s Respiratory Questionnaire (SGRQ) and the Seattle Obstructive Lung Disease Questionnaire (SOLQ). The SGRQ is a 50-item questionnaire developed at St George’s, University of London (SGUL) which includes three components/domains-Symptoms (frequency and severity), Activities (which exacerbate or are limited by the symptoms) and Impacts (social and psychological disturbances).^10^ The permissions for the use of the official Hindi translation were obtained from SGUL on August 1, 2019. The SOLQ is a 29-item questionnaire, which involves scoring across four dimensions, including three dimensions of health-physical function, emotional function and coping skills- and one of treatment satisfaction.^11^ A Hindi translation of SOLQ was prepared by the authors and was approved for use by the IEC. The two scales are scored differently; A subject with a high quality of life would likely score low on SGRQ and high on SOLQ. Likewise, a subject with more limitations would likely score high on SGRQ and low on SOLQ.

### 2.1 Sample size calculation

Taking the estimated persons with decreased quality of life to be 92% with precision of 7% and a confidence interval of 5%; the calculated sample size was 76.

### 2.2 Statistical analysis

Continuous data were presented as mean±standard deviation (SD), if normally distributed, and median [interquartile range (IQR)], if data were non-normal. Categorical variables were presented as frequency and percentages (n; %). Comparability of groups was analyzed by Chi-square test, Student’s t test or Mann-Whitney test as appropriate and results were deemed statistically significant if p<0.05. Statistical analysis was performed using StataCorp. 2017. Stata Statistical Software: Release 15. College Station, TX: StataCorp LP.

## 3. RESULTS

### 3.1 Patient characteristics

A total of 90 patients (mean age 40.4±11.6 years; 60 [66.7%] males) were enrolled in the study. The baseline clinical characteristics of the patients are tabulated in Table 1. Most (79 [87.8%]) of patients were non-smokers. Majority (30 [48.4%]) of the patients were undernourished (BMI <18.5 kg/m^2^).

**Table 1:** Baseline clinical and demographic characteristics of the patients (n= 90).

| Characteristics of patients | Mean±SD or N (%) |
| --- | --- |
| Age (years) (Mean±SD) | 40.4±11.6 |
| Age groups (years) | N (%) |
| 18-29 | 18 (20%) |
| 30-39 | 23 (25.6%) |
| 40-49 | 27 (30%) |
| 50-59 | 18 (20%) |
| ≥60 | 4 (4.4%) |
| Gender | N (%) |
| Male | 60 (66.7%) |
| Female | 30 (33.3%) |
| Smoking status | N (%) |
| Non-smoker | 79 (87.8%) |
| Active smoker | 4 (4.4%) |
| Reformed smoker | 7 (7.8%) |
| Occupation of patient | N (%) |
| Professional (White collar) | 2 (2.2%) |
| Semi-professional | 3 (3.3%) |
| Clerical, Shop owner | 10 (11.1%) |
| Skilled worker | 26 (28.9%) |
| Semi skilled worker | 12 (13.4%) |
| Unskilled worker | 3 (3.3%) |
| Unemployed | 34 (37.8%) |
| **Body Mass Index (kg/m<sup>2</sup>) (n=62)** | **N (%)** |
| <18.5 | 30 (48.4%) |
| 18.5-22.9 | 15 (24.2%) |
| 23-24.9 | 6 (9.7%) |
| ≥25 | 11 (17.7%) |
| **Comorbidities** | **N (%)** |
| Diabetes mellitus | 4 (4.5%) |
| Hypertension | 3 (3.3%) |
| No comorbidities | 83 (92.2%) |
| **On current treatment** | **N (%)** |
| Yes, inhaler treatment | 45 (50%) |
| Yes, hemostatic drugs | 13 (14.4%) |
| Yes, both | 1 (1.1%) |
| No | 31 (34.5%) |

### 3.2 Symptoms

Overall, 84 (93.3%(95% CI: 86.1%-97.5%)) patients were currently symptomatic (Table 2). The most common symptom was shortness of breath (67 [74.4%] patients) followed by cough (57 [63.3%] patients) and expectoration (48 [53.3%] patients). Most of the patients who experienced shortness of breath had a low grade dyspnea-mMRC grade 1 (28 [31.1%]) or grade 2 (25 [27.8%]).

**Table 2:** Symptoms reported by the study group (n=90)

| Symptom | N (%) | Mean (±SD) symptom duration (years) |
| --- | --- | --- |
| Asymptomatic | 6 (6.7%) |  |
| Shortness of breath | 67 (74.4%) | 6.0±6.5 |
| mMRC 0 (Dyspnea only with strenuous exercise) | 23 (25.6%) |  |
| mMRC 1 (Dyspnea when hurrying or walking up a slight hill) | 28 (31.1%) |  |
| mMRC 2 (Walks slower than people of the same age because of dyspnea or has to stop for breath when walking at own pace) | 25 (27.8%) |  |
| mMRC 3 (Stops for breath after walking 100 yards/91 m or after a few minutes) | 11 (12.2%) |  |
| mMRC 4 (Too dyspneic to leave house or breathless when dressing) | 3 (3.3%) |  |
| Cough | 57 (63.3%) | 7.1±6.7 |
| Expectoration | 48 (53.3%) | 6.4±6.9 |
| Chest pain | 20 (22.2%) | 5.2±5.2 |
| Hemoptysis | 29 (32.2%) | 6.4±8.2 |

Abbreviations: mMRC: Modified Medical Research Council scale grading of dyspnea; SD: Standard deviation.

### 3.3 Pulmonary function testing (n=63) and chest radiography (n=65)

A total of 63 patients could perform satisfactory pulmonary function tests (spirometry), out of which 61 (96.8%) were abnormal. Most (39 [61.9%]) of the patients had a mixed type of spirometric pattern as shown in Table 3. Out of 65 patients with chest radiographs available, 60 (92.3%) patients had abnormalities with 45 (69.2%) having bilateral chest radiographic abnormalities. The overall mean chest radiographic score was 6.7±4.2 (95% CI: 5.7-7.7).The patients with chest pain exhibited the maximum radiographic abnormalities scores (mean 8.4±4.4).

**Table 3:**
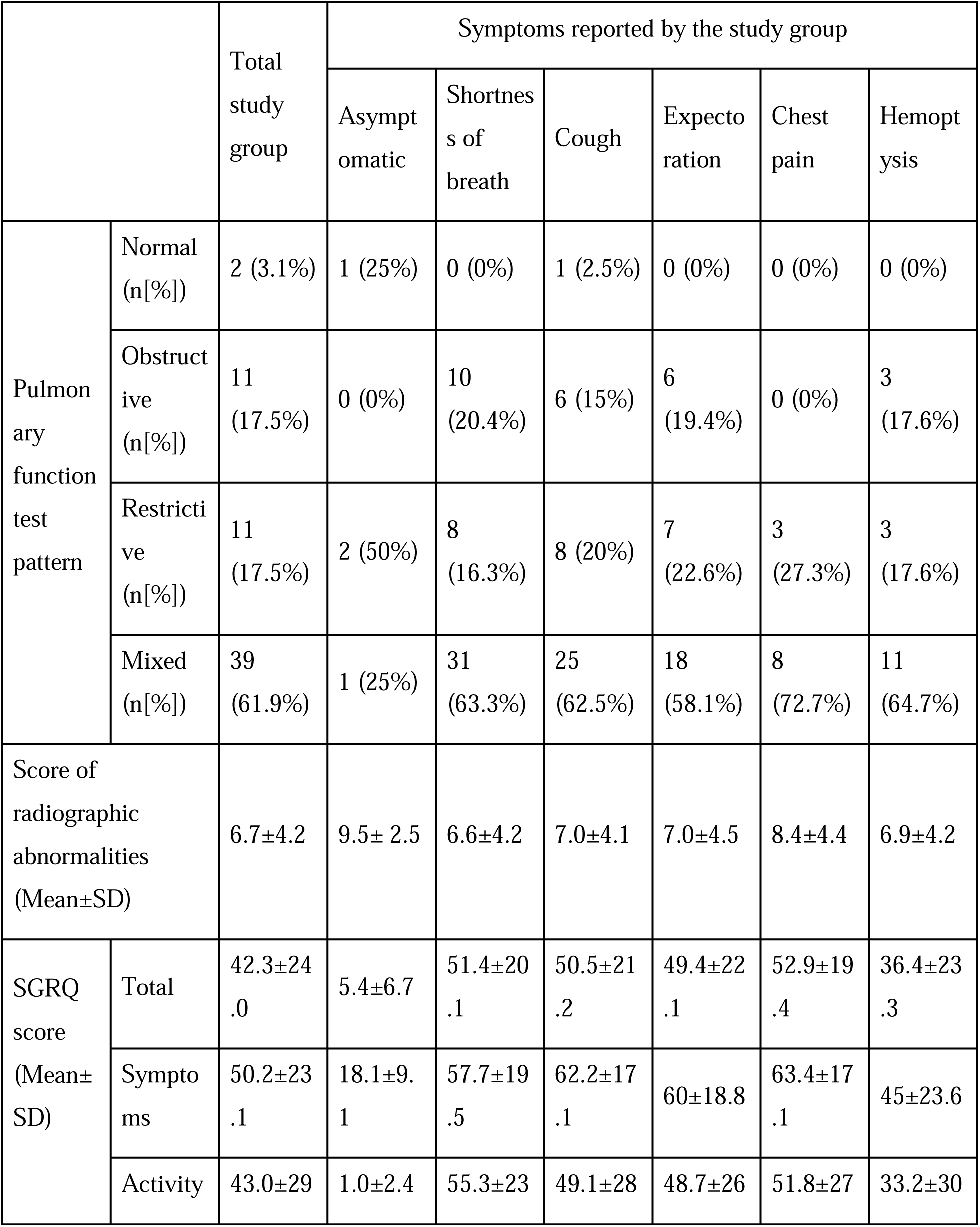
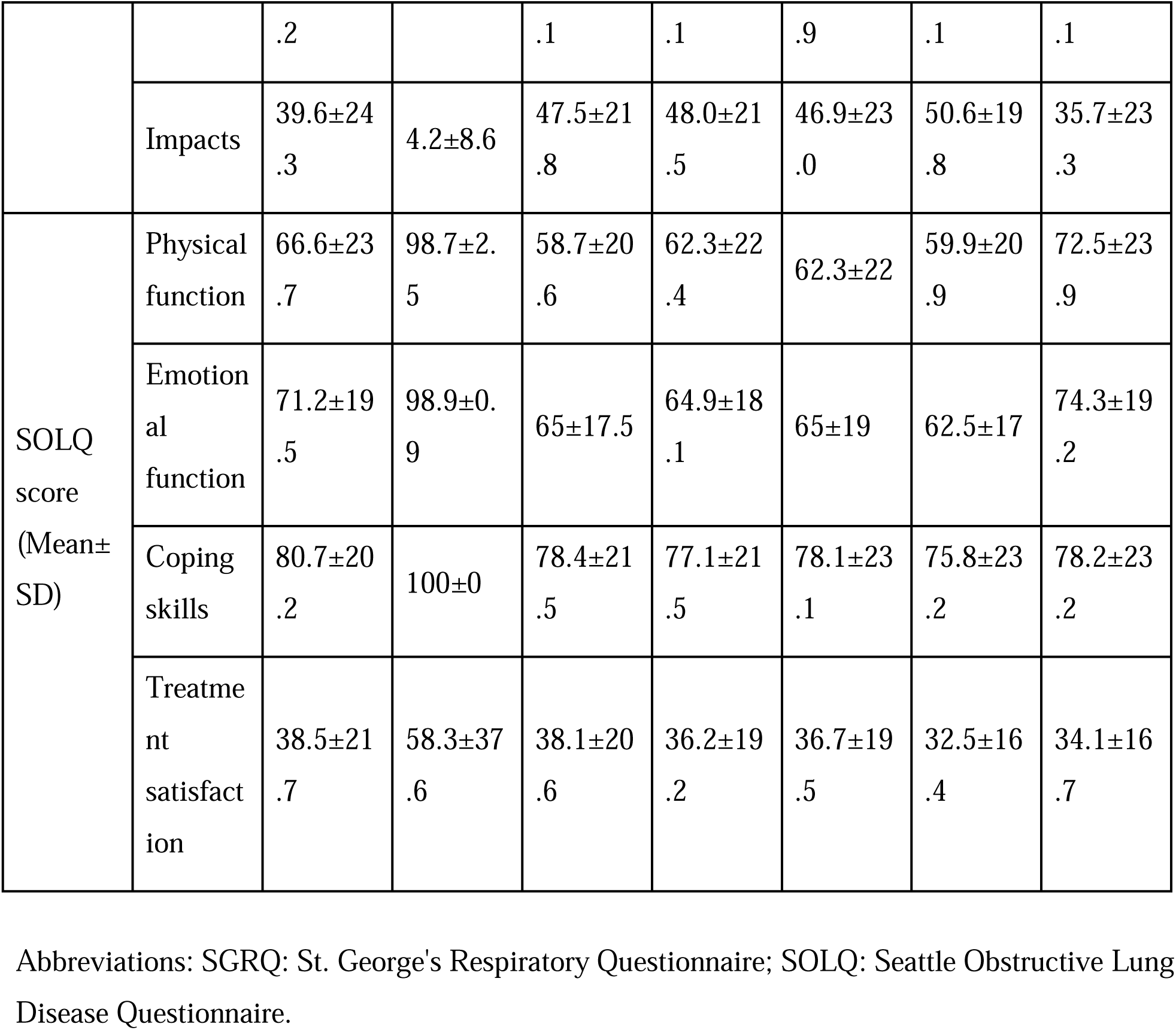
Association of symptoms reported by the patients with the imaging, pulmonary function test parameters, and the quality of life scores (n=90)

|  |  | Total study group | Symptoms reported by the study group |  |  |  |  |  |
| --- | --- | --- | --- | --- | --- | --- | --- | --- |
|  |  |  | Asymptomatic | Shortness of breath | Cough | Expectoration | Chest pain | Hemoptysis |
| Pulmonary function test pattern | Normal (n[%]) | 2 (3.1%) | 1 (25%) | 0 (0%) | 1 (2.5%) | 0 (0%) | 0 (0%) | 0 (0%) |
|  | Obstructive (n[%]) | 11 (17.5%) | 0 (0%) | 10 (20.4%) | 6 (15%) | 6 (19.4%) | 0 (0%) | 3 (17.6%) |
|  | Restrictive (n[%]) | 11 (17.5%) | 2 (50%) | 8 (16.3%) | 8 (20%) | 7 (22.6%) | 3 (27.3%) | 3 (17.6%) |
|  | Mixed (n[%]) | 39 (61.9%) | 1 (25%) | 31 (63.3%) | 25 (62.5%) | 18 (58.1%) | 8 (72.7%) | 11 (64.7%) |
| Score of radiographic abnormalities (Mean±SD) |  | 6.7±4.2 | 9.5± 2.5 | 6.6±4.2 | 7.0±4.1 | 7.0±4.5 | 8.4±4.4 | 6.9±4.2 |
| SGRQ score (Mean±SD) | Total | 42.3±24.0 | 5.4±6.7 | 51.4±20.1 | 50.5±21.2 | 49.4±22.1 | 52.9±19.4 | 36.4±23.3 |
|  | Symptoms | 50.2±23.1 | 18.1±9.1 | 57.7±19.5 | 62.2±17.1 | 60±18.8 | 63.4±17.1 | 45±23.6 |
|  | Activity | 43.0±29 | 1.0±2.4 | 55.3±23 | 49.1±28 | 48.7±26 | 51.8±27 | 33.2±30 |
|  |  | .2 |  | .1 | .1 | .9 | .1 | .1 |
|  | Impacts | 39.6±24<br>.3 | 4.2±8.6 | 47.5±21<br>.8 | 48.0±21<br>.5 | 46.9±23<br>.0 | 50.6±19<br>.8 | 35.7±23<br>.3 |
| SOLQ<br>score<br>(Mean±<br>SD) | Physical<br>function | 66.6±23<br>.7 | 98.7±2.<br>5 | 58.7±20<br>.6 | 62.3±22<br>.4 | 62.3±22 | 59.9±20<br>.9 | 72.5±23<br>.9 |
|  | Emotion<br>al<br>function | 71.2±19<br>.5 | 98.9±0.<br>9 | 65±17.5 | 64.9±18<br>.1 | 65±19 | 62.5±17 | 74.3±19<br>.2 |
|  | Coping<br>skills | 80.7±20<br>.2 | 100±0 | 78.4±21<br>.5 | 77.1±21<br>.5 | 78.1±23<br>.1 | 75.8±23<br>.2 | 78.2±23<br>.2 |
|  | Treatme<br>nt<br>satisfact<br>ion | 38.5±21<br>.7 | 58.3±37<br>.6 | 38.1±20<br>.6 | 36.2±19<br>.2 | 36.7±19<br>.5 | 32.5±16<br>.4 | 34.1±16<br>.7 |

Abbreviations: SGRQ: St. George's Respiratory Questionnaire; SOLQ: Seattle Obstructive Lung Disease Questionnaire.

### 3.4 Quality of life (QoL)

On assessment of QoL by SGRQ, the average score obtained was 42.3±24.0 (95% CI:37.3-47.3), with ‘Symptoms’ being the most affected domain on SGRQ (mean score 50.2±23.1). Patients complaining of chest pain had the worst QoL by SGRQ (52.9±19.4). On using SOLQ, ‘Treatment satisfaction’ (mean score 38.5±21.7, 95% CI:34-43) and ‘Physical function’ (mean score 66.6±23.7,95% CI:61.6-71.6) were the mainly affected parameters. The mean QoL scores in asymptomatic patients were, in general, lower on SGRQ and higher on SOLQ as compared to symptomatic patients, thus representing a higher QoL (Table 3). However, there was no correlation between the quality of life score of either of SOLQ or SGRQ with the chest radiographic score and spirometric pattern (Table 4).

**Table 4:** Correlation between different domains of the St. George’s Respiratory Questionnaire (SGRQ) and the Seattle Obstructive Lung Disease Questionnaire (SOLQ) and CXR score and Abnormal Spirometry {values are represented as regression coefficient [95% confidence interval]}

|  | SGRQ<br>total | Sympto<br>ms | Activity | Impact | SOLQ<br>total | SOLQ<br>PF | SOLQ<br>EF | SOLQ<br>TS | SOLQ<br>CS |
| --- | --- | --- | --- | --- | --- | --- | --- | --- | --- |
| CXR<br>RUQ<br>score | .01 [-<br>4.48,4.4<br>8] | -1.77 [-<br>6.04,2.5<br>0] | -.26 [-<br>5.67,5.1<br>5] | .64 [-<br>3.88,5.1<br>6] | -1.20 [-<br>4.58,2.1<br>7] | .45 [-<br>3.67,4.5<br>7] | -1.64 [-<br>5.17,1.8<br>8] | 4.74<br>[1.52,7.<br>96]* | -2.42 [-<br>6.13,1.3<br>0] |
| CXR<br>RLQ | 2.31 [-<br>2.10,6.7 | .05 [-<br>4.21,4.3 | 3.33 [-<br>1.97,8.6 | 2.40 [-<br>2.04,6.8 | -2.61 [-<br>6.02,.81 | -2.52 [-<br>6.70,1.6 | -2.92 [-<br>6.49,.64 | 6.11<br>[2.94,9. | -2.37 [-<br>6.19,1.4 |
| score | 2] | 1] | 3] | 4] | ] | 5] | ] | 28]* | 4] |
| CXR<br>LUQ<br>score | 1.58 [-<br>2.75,5.9<br>0] | 1.59 [-<br>2.55,5.7<br>3] | .99 [-<br>4.25,6.2<br>3] | 1.89 [-<br>2.47,6.2<br>5] | -.98 [-<br>4.29,2.3<br>4] | -.73 [-<br>4.76,3.3<br>1] | -2.08 [-<br>5.52,1.3<br>7] | 2.10 [-<br>1.23,5.4<br>2] | -.12 [-<br>3.82,3.5<br>6] |
| CXR<br>LLQ<br>score | 3.36 [-<br>1.62,1.3<br>4] | 3.82 [-<br>.92,8.57<br>] | 2.40 [-<br>3.68,8.4<br>7] | 3.80 [-<br>1.21,8.8<br>0] | -3.64 [-<br>7.51,.22<br>] | -3.19 [-<br>7.95,1.5<br>7] | -4.52 [-<br>8.52,-<br>.51]* | 1.92 [-<br>2.08,5.9<br>2] | -3.22 [-<br>7.55,1.1<br>1] |
| CXR<br>total<br>score | .75 [-<br>.35,1.85<br>] | .47 [-<br>.60,1.53<br>] | .81 [-<br>.53,2.15<br>] | .80 [-<br>.31,1.91<br>] | -.60 [-<br>1.44,.23<br>] | -.39 [-<br>1.47,.69<br>] | -.87 [-<br>1.74,.00<br>]* | .38 [-<br>.60,1.37<br>] | -.55 [-<br>1.47,.36<br>] |
| Abnorm<br>al<br>Spirome<br>try | 2.94 [-<br>7.42,13.<br>31] | 2.39 [-<br>7.59,12.<br>36] | 4.45 [-<br>8.13,17.<br>02] | 2.12 [-<br>8.36,12.<br>61] | -.09 [-<br>8.05,7.8<br>6] | -2.01 [-<br>12.22,8.<br>21] | .69 [-<br>7.74,9.1<br>2] | -3.01 [-<br>12.35,6.<br>32] | 1.04 [-<br>7.70,9.7<br>8] |

\*p<0.05 , Abbreviations: PF: Physical function; EF: Emotional function; CS: Coping skills; TS: Treatment satisfaction; CXR: Chest X-ray; RUQ: Right Upper Quadrant; RLQ: Right Lower Quadrant; LUQ: Left Upper Quadrant; LLQ: Left Lower Quadrant

On comparing our two QoL instruments, the scores obtained on SGRQ negatively correlated with the scores on SOLQ with most being statistically significant (p<0.05), as shown in Table 5.

**Table 5:**
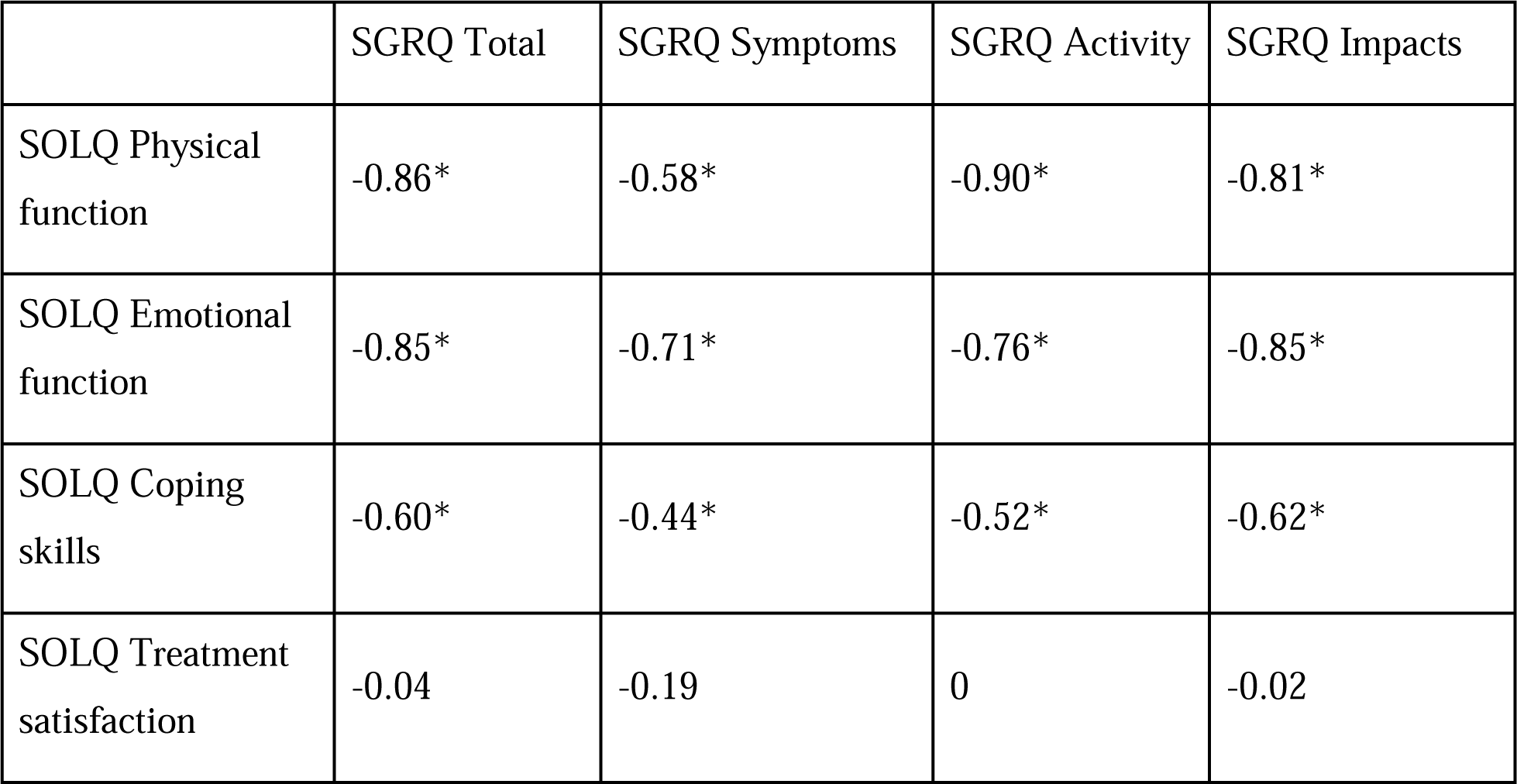
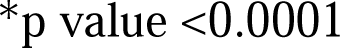
Pearson correlation coefficients between different domains of the St. George’s Respiratory Questionnaire (SGRQ) and the Seattle Obstructive Lung Disease Questionnaire (SOLQ).

|  | SGRQ Total | SGRQ Symptoms | SGRQ Activity | SGRQ Impacts |
| --- | --- | --- | --- | --- |
| SOLQ Physical function | -0.86* | -0.58* | -0.90* | -0.81* |
| SOLQ Emotional function | -0.85* | -0.71* | -0.76* | -0.85* |
| SOLQ Coping skills | -0.60* | -0.44* | -0.52* | -0.62* |
| SOLQ Treatment satisfaction | -0.04 | -0.19 | 0 | -0.02 |

\*p value <0.0001

## 4. DISCUSSION

The present study shows that patients with post-TB sequelae have significant symptoms, radiological abnormalities, spirometry derangements, and impaired QoL. Amongst these patients, shortness of breath and cough were the most common symptoms with expectoration, chest pain, and hemoptysis being the other significant symptoms. Majority of patients had abnormal spirometry, of which mixed pattern was the most common. Bilateral chest radiographic abnormalities were seen in 69.2% of our patients. There were significant differences in the QoL parameters between asymptomatic and symptomatic patients. Patients with chest pain had the most severely affected QoL by SGRQ. There were statistically significant inter-questionnaire correlations between the QoL parameters measured by SGRQ and SOLQ.

In our study cohort, 93.3% patients were symptomatic with shortness of breath and cough being the most common symptoms. Similarly, high prevalence of respiratory symptoms (78-95%) has been observed in previous studies.^8,13–15^ In our study, the most frequent presenting symptom was shortness of breath, while other studies have reported cough (Akkara et al)^14^ and hemoptysis (Panda et al)^13^ to be the most common symptom.

Despite chemotherapy, widespread tissue destruction from TB can be associated with high mortality and morbidity. Healing of the affected tissue may be accompanied by extensive fibrosis, traction bronchiectasis, bronchostenosis, and abnormalities on PFT.^8,16^ The prevalence of post-TB bronchiectasis has been reported to be 35-86% in a recent systematic review.^17^ Additionally, extensive fibrosis may result in stiffening of the lung parenchyma leading to restrictive lung disease, where patients may have cough, chest pain and shortness of breath. Inflammation-induced narrowing of airways may result in airflow obstruction, which may present with dyspnea and exercise intolerance.^2,18^ Bronchiectasis and erosion of blood vessels by cavitation may result in hemoptysis.^16^ It is likely that these patterns of healing are the result of the variation in the effector responses by the host immune system.^2,16^

In our study, mixed ventilatory defect was the most common type of pattern on PFT (61.9%). Previous studies have shown obstructive,^14^ restrictive,^7,13,19^ or mixed pattern^8,15,20^ to be the most common PFT abnormality.

We observed 92.3% of our patients to have abnormal chest X-ray while 69.2% had bilateral radiographic abnormalities. In previous studies, abnormal chest radiographs have been observed in 71.9-100% of patients.^8,15,19,21^ It has been reported that extensive residual lung lesions may predict tissue destruction and an increased predisposition to opportunistic infections, thus leading to permanent disability and a reduced QoL.^8^ Significant correlations have also been shown between the extent of radiological abnormalities on chest computed tomography and dyspnea.^12^

In our study, QoL was observed to be adversely impaired via both our QoL instruments. Despite there being several studies on QoL in patients with active TB or those undergoing treatment,^22–24^ very few studies have focused on QoL in post-TB patients, especially focusing on those with long term sequelae. Muniyandi et al,^25^ using the SF-36 questionnaire, reported that the QoL of TB patients one year after completion of anti-tubercular therapy (ATT) was normal. However, data from the US general population was used as the comparator for Indian patients. Singla et al^8^ reported an impaired QoL using the SOLQ in 46 patients with MDR-TB after completion of ATT. Gandhi et al^19^ recruited patients who had recently completed ATT and reported that patients with impairment on PFT had lower scores on SOLQ across all four domains. Similar to our study, ‘Treatment satisfaction’ domain was the most affected in the study by Gandhi et al, while Singla et al observed ‘Physical function’ to be the most affected.

TB sequelae are likely to result in patients with delayed diagnosis, extensive disease, and long/repeated treatments.^4^ As pharmacological therapies have a limited impact, these patients may benefit from early initiation of other supportive interventions which are likely to have a positive impact. These include pulmonary rehabilitation, smoking cessation, vaccination against secondary infections, and psychological and nutritional support.^4,26^ Our study highlights the need for early diagnosis and initiation of appropriate treatment for patients with active TB as well as those with post-TB sequelae. Long term sequelae must always be considered as a differential in symptomatic patients with a history of TB.

The strength of our study is the large consecutive sample of patients along with inclusion of patients with a history of TB even in the distant past. It reflects the quality of life of tubercular sequelae patients, with tuberculosis being one of the most important disease burdens in India. Two well accepted, robust, validated QoL instruments were used in our study for all patients with each of them being administered by the authors, instead of being self-administered. These questionnaires thus can be translated and used in other settings as well. A limitation of our study is that it’s a single center cross-sectional study and there is a lack of follow-up data for these patients. There is potential selection bias as only moderate and severe cases report to clinic, missing out on the mild cases.Thus, our study may not reflect the full spectrum of disease in post-TB patients as only the patients presenting to the chest clinic were included, which may limit the generalisability of our results for the TB sequelae patients in general.

## 5. CONCLUSION

In conclusion, patients with a history of pulmonary TB end up with post-TB sequelae and have respiratory symptoms. Most of our study patients had radiographic abnormalities and abnormal PFT with mixed pattern being the most common. QoL, as measured by SGRQ and SOLQ, was significantly impaired in these patients. Significant correlations were seen in the measurements between our two QoL instruments. Long term sequelae must always be considered as a differential in symptomatic patients with a history of TB. Further studies are needed to evaluate the role of other factors that can affect the QoL.

## Data Availability

Data is available on reasonable request.

## Acknowledgements

We thank all the participating volunteers for their efforts and contributions.

## REFERENCES

1. Allwood BW, van der Zalm MM, Amaral AFS, Byrne A, Datta S, Egere U, et al. Post-tuberculosis lung health: perspectives from the First International Symposium. Int J Tuberc Lung Dis. 2020 Aug 1;24(8):820–8.

2. Ravimohan S, Kornfeld H, Weissman D, Bisson GP. Tuberculosis and lung damage: from epidemiology to pathophysiology. Eur Respir Rev. 2018 Mar 31;27(147).

3. Romanowski K, Baumann B, Basham CA, Ahmad Khan F, Fox GJ, Johnston JC. Long-term all-cause mortality in people treated for tuberculosis: a systematic review and meta-analysis. Lancet Infect Dis. 2019 Oct;19(10):1129–37.

4. Visca D, Centis R, Munoz-Torrico M, Pontali E. Post-tuberculosis sequelae: the need to look beyond treatment outcome. Int J Tuberc Lung Dis. 2020 Aug 1;24(8):761–2.

5. von Elm E, Altman DG, Egger M, Pocock SJ, Gøtzsche PC, Vandenbroucke JP, et al. The Strengthening the Reporting of Observational Studies in Epidemiology (STROBE) statement: guidelines for reporting observational studies. Ann Intern Med. 2007 Oct 16;147(8):573–7.

6. Wani RT. Socioeconomic status scales-modified Kuppuswamy and Udai Pareekh’s scale updated for 2019. J Family Med Prim Care. 2019 Jun;8(6):1846–9.

7. de Vallière S, Barker RD. Residual lung damage after completion of treatment for multidrug-resistant tuberculosis. Int J Tuberc Lung Dis. 2004 Jun;8(6):767–71.

8. Singla R, Mallick M, Mrigpuri P, Singla N, Gupta A. Sequelae of pulmonary multidrug-resistant tuberculosis at the completion of treatment. Lung India. 2018 Feb;35(1):4–8.

9. Báez-Saldaña R, López-Arteaga Y, Bizarrón-Muro A, Ferreira-Guerrero E, Ferreyra-Reyes L, Delgado-Sánchez G, et al. A novel scoring system to measure radiographic abnormalities and related spirometric values in cured pulmonary tuberculosis. PLoS One. 2013;8(11):e78926.

10. Jones PW, Quirk FH, Baveystock CM, Littlejohns P. A self-complete measure of health status for chronic airflow limitation. The St. George’s Respiratory Questionnaire. Am Rev Respir Dis. 1992 Jun;145(6):1321–7.

11. Tu SP, McDonell MB, Spertus JA, Steele BG, Fihn SD. A new self-administered questionnaire to monitor health-related quality of life in patients with COPD. Ambulatory Care Quality Improvement Project (ACQUIP) Investigators. Chest. 1997 Sep;112(3):614–22.

12. Sathiyamoorthy R, Kalaivani M, Aggarwal P, Gupta SK. Prevalence of pulmonary tuberculosis in India: A systematic review and meta-analysis. Lung India Off Organ Indian Chest Soc. 2020 Feb;37(1):45–52.

13. Panda A, Bhalla AS, Sharma R, Mohan A, Sreenivas V, Kalaimannan U, et al. Correlation of chest computed tomography findings with dyspnea and lung functions in post-tubercular sequelae. Lung India. 2016 Dec;33(6):592–9.

14. Akkara SA, Shah AD, Adalja M, Akkara AG, Rathi A, Shah DN. Pulmonary tuberculosis: the day after. Int J Tuberc Lung Dis. 2013 Jun;17(6):810–3.

15. Singla N, Singla R, Fernandes S, Behera D. Post treatment sequelae of multi-drug resistant tuberculosis patients. Indian J Tuberc. 2009 Oct;56(4):206–12.

16. Dheda K, Booth H, Huggett JF, Johnson MA, Zumla A, Rook GAW. Lung remodeling in pulmonary tuberculosis. J Infect Dis. 2005 Oct 1;192(7):1201–9.

17. Meghji J, Simpson H, Squire SB, Mortimer K. A Systematic Review of the Prevalence and Pattern of Imaging Defined Post-TB Lung Disease. PLoS One. 2016;11(8):e0161176.

18. Vogelmeier CF, Criner GJ, Martinez FJ, Anzueto A, Barnes PJ, Bourbeau J, et al. Global Strategy for the Diagnosis, Management, and Prevention of Chronic Obstructive Lung Disease 2017 Report: GOLD Executive Summary. Eur Respir J. 2017 Mar;49(3).

19. Gandhi K, Gupta S, Singla R. Risk factors associated with development of pulmonary impairment after tuberculosis. Indian J Tuberc. 2016 Jan;63(1):34–8.

20. Ramos LMM, Sulmonett N, Ferreira CS, Henriques JF, de Miranda SS. Functional profile of patients with tuberculosis sequelae in a university hospital. J Bras Pneumol. 2006 Feb;32(1):43–7.

21. Kumar Rai D, Kumar R. Identification of risk factors for radiological sequelae in patients treated for pulmonary tuberculosis: Prospective observational cohort study. Indian J Tuberc. 2020 Oct;67(4):534–8.

22. Aggarwal AN, Gupta D, Janmeja AK, Jindal SK. Assessment of health-related quality of life in patients with pulmonary tuberculosis under programme conditions. Int J Tuberc Lung Dis. 2013 Jul;17(7):947–53.

23. Atif M, Sulaiman SAS, Shafie AA, Asif M, Sarfraz MK, Low HC, et al. Impact of tuberculosis treatment on health-related quality of life of pulmonary tuberculosis patients: a follow-up study. Health Qual Life Outcomes. 2014 Feb 14;12:19.

24. Maguire GP, Anstey NM, Ardian M, Waramori G, Tjitra E, Kenangalem E, et al. Pulmonary tuberculosis, impaired lung function, disability and quality of life in a high-burden setting. Int J Tuberc Lung Dis. 2009 Dec;13(12):1500–6.

25. Muniyandi M, Rajeswari R, Balasubramanian R, Nirupa C, Gopi PG, Jaggarajamma K, et al. Evaluation of post-treatment health-related quality of life (HRQoL) among tuberculosis patients. Int J Tuberc Lung Dis. 2007 Aug;11(8):887–92.

26. Tiberi S, Torrico MM, Rahman A, Krutikov M, Visca D, Silva DR, et al. Managing severe tuberculosis and its sequelae: from intensive care to surgery and rehabilitation. J Bras Pneumol [Internet]. 2019 [cited 2021 Mar 17];45(2). Available from: https://www.ncbi.nlm.nih.gov/pmc/articles/PMC6733754/

